# Learning Where to Look for COVID-19 Growth: Multivariate Analysis of COVID-19 Cases Over Time using Explainable Convolution-LSTM

**DOI:** 10.1101/2021.02.13.21251683

**Authors:** Novanto Yudistira, Sutiman Bambang Sumitro, Alberth Nahas, Nelly Florida Riama

## Abstract

Determinant factors which contribute to the prediction should take into account multivariate analysis for capturing coarse-to-fine contextual information. From the preliminary descriptive analysis, it shows that environmental factor such as UV (ultraviolet) is one of the essential factors that should be considered to observe the COVID-19 epidemic drivers, During summer, UV can inactivate viruses that live in the air and on the surface of the objects especially at noon in tropical or subtropical countries. However, it may not be significant in closed spaces like workspace and areas with the intensive human-to-human transmission, especially in densely populated areas. Different COVID-19 pandemic growth patterns in northern subtropical, southern subtropical and tropical countries occur over time. Moreover, there are education, government, morphological, health, economic, and behavioral factors contributing to the growth of COVID-19. Multivariate analysis via visual attribution of explainable Convolution-LSTM is utilized to see high contributing factors responsible for the growth of daily COVID-19 cases. For future works, data to be analyzed should be more detailed in terms of the region and the period where the time-series sample is acquired. The explainable Convolution-LSTM code is available here: https://github.com/cbasemaster/time-series-attribution

## 1. Introduction

The Wuhan Municipal Health Commission first detected the 2019 Coronavirus Disease (COVID-19) in Hubei Province, China, and early information about the outbreak has been sent to the World Health Organization (WHO) [1] [2]. As the number of people exposed to COVID-19 increases, the disease’s ability to spread in the community has rapidly improved. The rapid growth involves evidence of person-to-person transmission, indicating that COVID-19 is highly contagious. COVID-19 can also live actively in the air and on the ground [3] [4]. Environmental factors are affecting the success of airborne viruses spreading among susceptible hosts [5]. These forms of transmission may cause a pandemic. A pandemic can cause severe activity and death in a wide geographic area [6]. To withstand the spread of COVID-19, a natural factor that is rarely discussed is ultraviolet (UV). Many UV papers prove that it can inactivate viruses [7] [8], especially ultraviolet rays from sunlight [9]. An example of virus inactivation by ultraviolet light is ultraviolet-C radiation for virus inactivation [8]. Even though ultraviolet rays can inactivate the virus, it will not be evident if the pollution level is high [10]. Please note that smoke particles will weaken ultraviolet light’s ability to exist in the air [11]. Efforts to reduce the spread of COVID-19 can also reduce carbon emissions by reducing the intensity of travel worldwide [12], which is good for the environment. Moreover, vaccine development is not sufficient, and it takes a long time to discover [13]. Therefore, urgent, large-scale, and natural immunity is needed. Some technologies have been developed by using UV light [14] [15]. Based on the above evidence, we investigate how UV rays dynamically affect the spread of COVID-19 based on geographic location, pollution levels, and human activities. Multivariate time series data analysis is a better choice for analyzing the growth of the COVID-19 pandemic because it has interdependence among multiple factors over time. The classification of multivariate time series is also an emerging hot topic in machine learning [19]. Deep neural network (DNN) can capture information from big data, thus it is the best candidate to perform classification tasks [20] [21]. The ability of DNN to generate meaningful feature representations in the learning process has attracted attention in the machine learning and data science circles. In this study, we use interpretable DNN forecasts to perform multivariate time series data analysis. This explanation helps to find critical joint characteristics to predict daily cases of COVID-19 over a period of time. One study using interpretable DNN is Roy Assaf et al.’s multivariate multi-factory PV energy prediction, [24] which uses a two-stage convolutional neural network (CNN).

## 2. Materials and methods

### 2.1. Data sets

This multidimensional study uses 3 data sets of COVID-19 growth and its attribution, UVs, and people mobility data. The time-series data was taken from 2020-03-22 until 2020-09-11. The selected countries are located in tropical, northern subtropical, and southern subtropical regions. Data sets of worlds confirmed COVID-19, UV index, pollution, and people mobility time series were taken from Ourworldindata[1], Tropospheric Emission Monitoring Internet Service (TEMIS)[2] [16], and Google Mobility[3], respectively. Specific data, like UV index and pollution in Jakarta, Indonesia, were taken from IndonesiaMeteorology, Climatology, and Geophysical Agency (BMKG)[4]. Confirmed, recovered, and death cases of COVID-19 data in Jakarta have been obtained from the Indonesia Ministry of Health[5].

Table 1 shows 59 factors used in this COVID-19 growth multivariate time-series analysis ranging from environmental, social, government, economical to behavioral factors. The clear-sky UV index (UVIEF) is a measure for the effective UV irradiance (1 unit equals 25 mW/m2) reaching the Earth’s surface. The UV dose is the effective UV irradiance (given in kJ/m2), reaching the Earth’s surface integrated over the day and taking the UV radiation’s attenuation due to clouds. The cloud data is compiled from the geostationary Meteosat Second Generation (MSG) observations. The UV dose is computed for three different action spectra, i.e., for three other health effects: erythema (sunburn) of the skin (UVDEF), vitamin-D production in the skin (UVDVF), and DNA-damage (UVDDF).

**Table 1.**
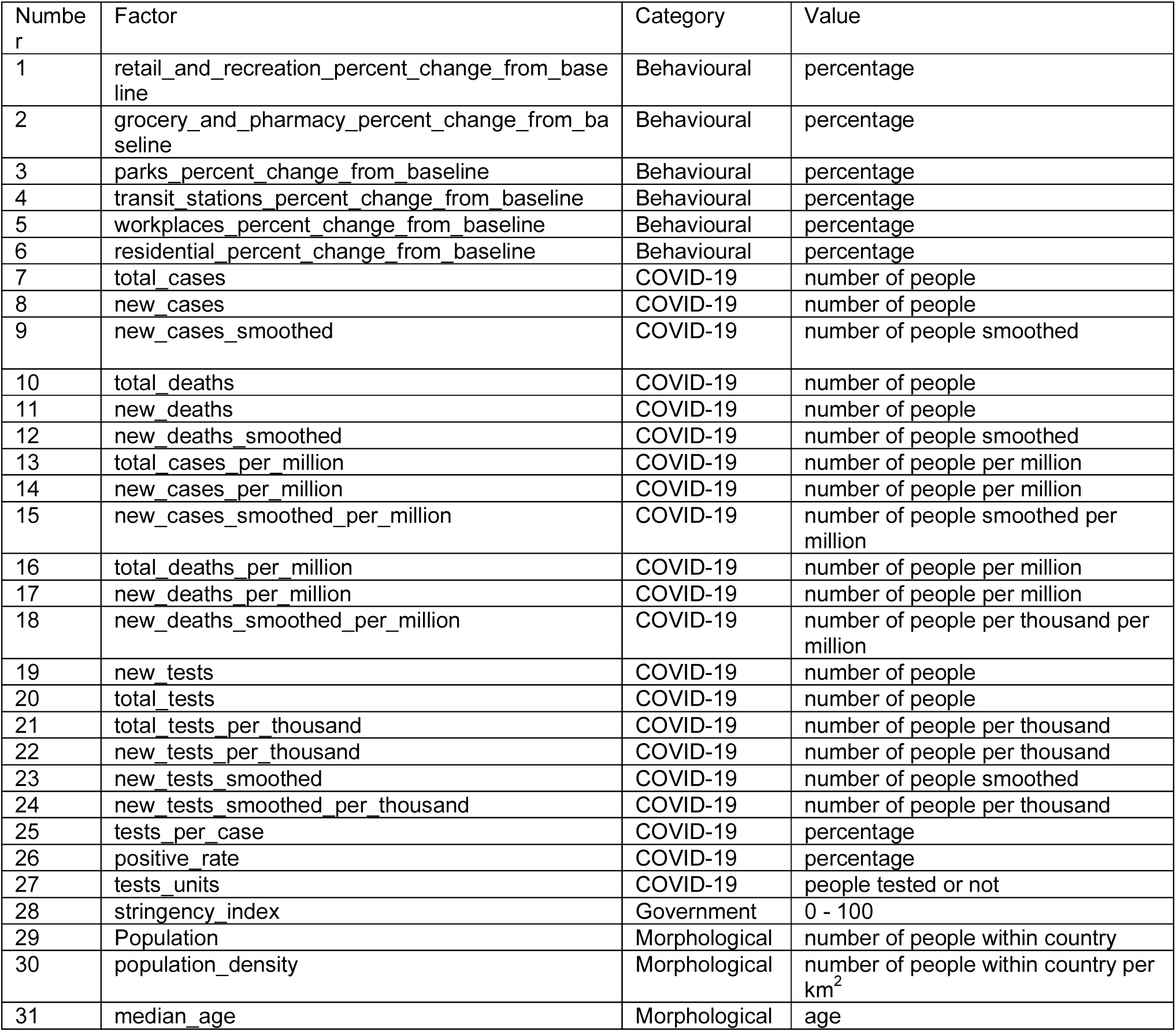
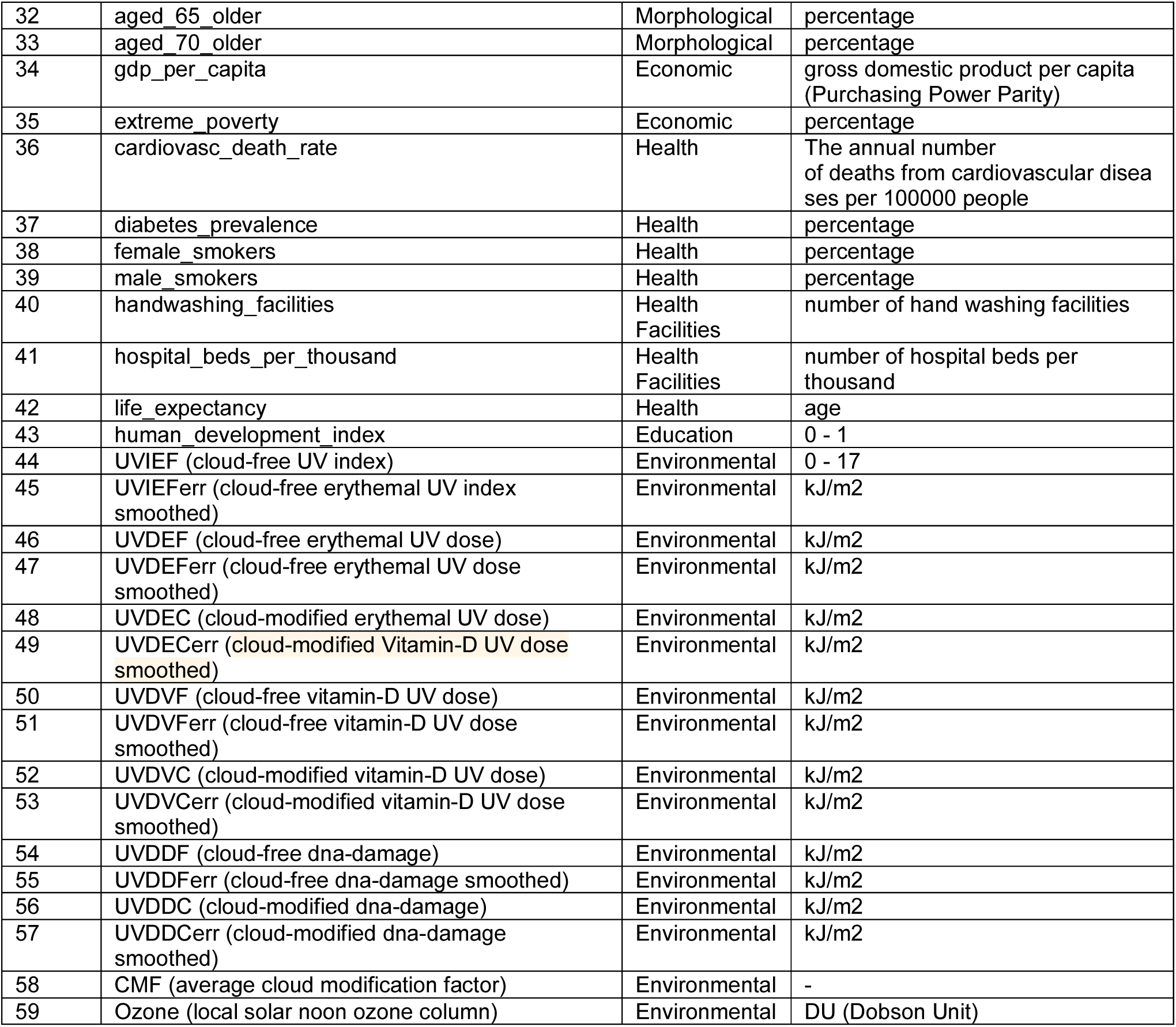
59 Factors used in this research ranging from environmental, government, economical, to behavioural factors during 2020-03-22 to 2020-09-11.

| Number | Factor | Category | Value |
| --- | --- | --- | --- |
| 1 | retail_and_recreation_percent_change_from_baseline | Behavioural | percentage |
| 2 | grocery_and_pharmacy_percent_change_from_baseline | Behavioural | percentage |
| 3 | parks_percent_change_from_baseline | Behavioural | percentage |
| 4 | transit_stations_percent_change_from_baseline | Behavioural | percentage |
| 5 | workplaces_percent_change_from_baseline | Behavioural | percentage |
| 6 | residential_percent_change_from_baseline | Behavioural | percentage |
| 7 | total_cases | COVID-19 | number of people |
| 8 | new_cases | COVID-19 | number of people |
| 9 | new_cases_smoothed | COVID-19 | number of people smoothed |
| 10 | total_deaths | COVID-19 | number of people |
| 11 | new_deaths | COVID-19 | number of people |
| 12 | new_deaths_smoothed | COVID-19 | number of people smoothed |
| 13 | total_cases_per_million | COVID-19 | number of people per million |
| 14 | new_cases_per_million | COVID-19 | number of people per million |
| 15 | new_cases_smoothed_per_million | COVID-19 | number of people smoothed per million |
| 16 | total_deaths_per_million | COVID-19 | number of people per million |
| 17 | new_deaths_per_million | COVID-19 | number of people per million |
| 18 | new_deaths_smoothed_per_million | COVID-19 | number of people per thousand per million |
| 19 | new_tests | COVID-19 | number of people |
| 20 | total_tests | COVID-19 | number of people |
| 21 | total_tests_per_thousand | COVID-19 | number of people per thousand |
| 22 | new_tests_per_thousand | COVID-19 | number of people per thousand |
| 23 | new_tests_smoothed | COVID-19 | number of people smoothed |
| 24 | new_tests_smoothed_per_thousand | COVID-19 | number of people per thousand |
| 25 | tests_per_case | COVID-19 | percentage |
| 26 | positive_rate | COVID-19 | percentage |
| 27 | tests_units | COVID-19 | people tested or not |
| 28 | stringency_index | Government | 0 - 100 |
| 29 | Population | Morphological | number of people within country |
| 30 | population_density | Morphological | number of people within country per km<sup>2</sup> |
| 31 | median_age | Morphological | age |
| 32 | aged_65_older | Morphological | percentage |
| 33 | aged_70_older | Morphological | percentage |
| 34 | gdp_per_capita | Economic | gross domestic product per capita (Purchasing Power Parity) |
| 35 | extreme_poverty | Economic | percentage |
| 36 | cardiovasc_death_rate | Health | The annual number of deaths from cardiovascular diseases per 100000 people |
| 37 | diabetes_prevalence | Health | percentage |
| 38 | female_smokers | Health | percentage |
| 39 | male_smokers | Health | percentage |
| 40 | handwashing_facilities | Health Facilities | number of hand washing facilities |
| 41 | hospital_beds_per_thousand | Health Facilities | number of hospital beds per thousand |
| 42 | life_expectancy | Health | age |
| 43 | human_development_index | Education | 0 - 1 |
| 44 | UVIEF (cloud-free UV index) | Environmental | 0 - 17 |
| 45 | UVIEFerr (cloud-free erythemal UV index smoothed) | Environmental | kJ/m2 |
| 46 | UVDEF (cloud-free erythemal UV dose) | Environmental | kJ/m2 |
| 47 | UVDEFerr (cloud-free erythemal UV dose smoothed) | Environmental | kJ/m2 |
| 48 | UVDEC (cloud-modified erythemal UV dose) | Environmental | kJ/m2 |
| 49 | UVDECerr (cloud-modified Vitamin-D UV dose smoothed) | Environmental | kJ/m2 |
| 50 | UVDVF (cloud-free vitamin-D UV dose) | Environmental | kJ/m2 |
| 51 | UVDVFerr (cloud-free vitamin-D UV dose smoothed) | Environmental | kJ/m2 |
| 52 | UVDVC (cloud-modified vitamin-D UV dose) | Environmental | kJ/m2 |
| 53 | UVDVCerr (cloud-modified vitamin-D UV dose smoothed) | Environmental | kJ/m2 |
| 54 | UVDDF (cloud-free dna-damage) | Environmental | kJ/m2 |
| 55 | UVDDFerr (cloud-free dna-damage smoothed) | Environmental | kJ/m2 |
| 56 | UVDDC (cloud-modified dna-damage) | Environmental | kJ/m2 |
| 57 | UVDDCerr (cloud-modified dna-damage smoothed) | Environmental | kJ/m2 |
| 58 | CMF (average cloud modification factor) | Environmental | - |
| 59 | Ozone (local solar noon ozone column) | Environmental | DU (Dobson Unit) |

Government stringency is related to the measurement provided by the Oxford COVID-19 Government Reaction Tracker [26]. The Tracker includes 100 Oxford community individuals who have ceaselessly upgraded a database of 17 parameters of government response. These parameters look at control approaches such as school and working environment closings, open occasions, open transport, and stay-at-home policies. The Stringency Record could be a number from 0 to 100 that reflects these indicators. The higher score shows a better level of stringency. Stringency list gives a picture of the policies at which any nation implemented its most grounded measures. Some countries have had their deaths continue to flatten only as they have hit their hardest stringency, such as Italy, Spain, or France. As China took harder than initial stringency, its death curve was flattened.

The aforementioned data set references can be accessed to investigate the detailed definition of each factor. There are 55 countries at various scales of geographical area and population trained on DNNs to reveal its explanations. Note that all factors are normalized into the 0-1 range before feeding into DNNs.

### 2.2. UV Index Dynamics as Environmental Factors

The UV index (UVIEF) is derived from the measured solar radiation in the UV spectra that arrives on the surface. It is calculated by considering the proportional contribution of UV-A and UV-B, two of the three-wavelength based types of UV radiation. UV-A is characterized as the UV radiation of which the wavelength ranges from 280-315 nm, while the wavelength of UV-B is between 315 nm and 400 nm. UV spectra are captured by the Global Atmosphere Watch (GAW) station. In this study, the daily mean UV index (0-17) and daily mean Ozone (Dobson Unit) are considered as parameters involved in the correlation analysis. The depletion of the protective stratospheric ozone layer due to chlorofluorocarbons (CFCs) and halons has increased UV radiation. The UV index is measured under the assumption of a clear sky without any barriers such as clouds. The trade-offs between 3 geographical locations (northern subtropical, tropical, and southern subtropical areas) are investigated in terms of how UV index and COVID-19 are related over time. The selected GAWs are located in Argentina, Australia, Chile, Brazil (Sao Paulo) for southern subtropical countries. For tropical countries, GAWs of India, Saudi Arabia, Thailand are selected as representative countries. Finally, GAWs of Germany, Italy, Japan, Russia, Spain, Taiwan are selected to represent northern subtropical countries. The duration UV index time series is from 2020/01/22 to 2020/07/20. The time-series data are then transformed into a weekly mean series to capture the bigger picture of dynamics.

Fig.1a, 1b, and 1c show pandemic growth in tropical (green), northern subtropical (blue), and southern subtropical (red) countries for confirmed, recovered, and death cases, respectively. Note that the number of confirmed cases, recovered, and deaths are normalized across all countries. Since the end of March, the blue countries grew over time and faster than the green and red countries. The green countries grew sharper and faster than red countries, even though there were cross points in the middle of growth. The overtaking points indicate a growth pace that is becoming slower than the other, and vice versa. This dynamical pace happened between two adjacent groups, either blue with green or green with red. Blue countries were starting to converge, and conversely, red countries were starting to emerge both at the beginning of May even though outlier countries exist. These phenomena possibly can be explained in fig.1d where daily mean UV index in red countries were monotonically decreasing as the winter comes and the opposite for blue countries. It can be suggested that there is an indication that COVID-19 is a seasonal pandemic depending on geographical locations.

**Figure 1.**
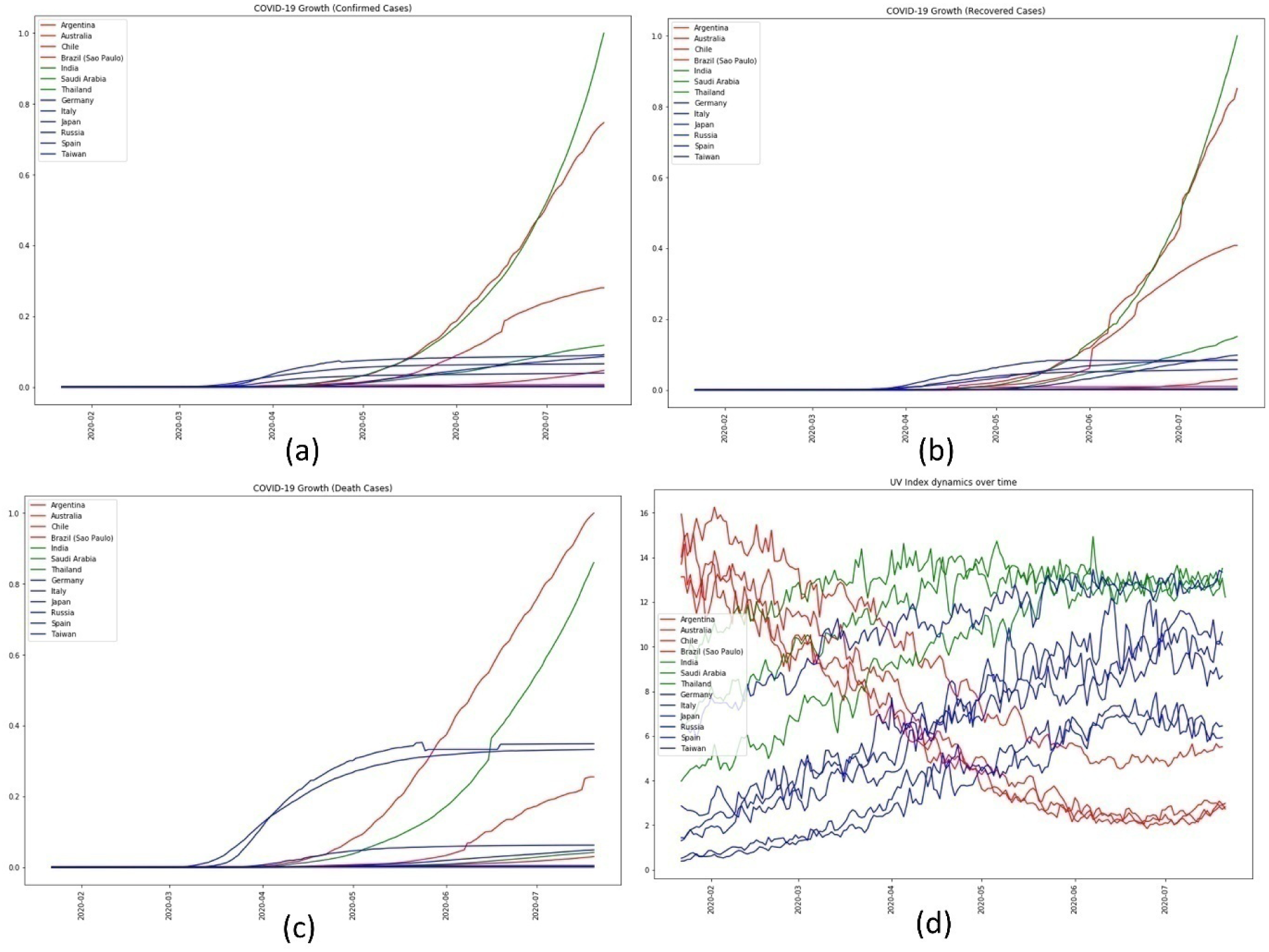
(a) The growth of cumulative confirmed cases in northern subtropical (blue), tropical (green), and southern subtropical (red) countries (b) The growth of cumulative recovered cases in northern subtropical (blue), tropical (green), and southern subtropical (red) countries (c) The growth of cumulative death cases in northern subtropical (blue), tropical (green), and southern subtropical (red) countries (d) Daily mean UV Index dynamics over time of northern subtropical countries (blue), tropical (green) and southern subtropical (red)

### 2.3. Human Mobility Dynamics as Behavioural Factors

The key drivers to be used in human mobility analysis are community activities dynamics during a pandemic depending on geographical locations (focusing on tropical countries). After the first outbreak, human mobility has changed from before pandemic due to lockdown or outdoor activity restriction from the government. To see the effect of these restrictions, we investigate activities dynamics relative to COVID-19 growth. To realize this, Google Mobility data that provide six different activities are utilized. Those activities are grocery and pharmacy, workplaces, transit stations, retail and recreation, residential, and parks percent change from baseline. To see the effect of reducing activities intensity, we analyse the time-lagged correlation between activities dynamics and COVID-19 growth. It means that the impact of activities reductions on COVID-19 growth patterns after several days is temporally investigated. The countries to be investigated in this study are India, Brazil, Malaysia, Saudi Arabia, Indonesia, Thailand (tropical countries).

Fig. 2a shows that human mobility in Brazil reached the lowest activities percent change to baseline in the middle of April 2020 and then gradually increased its percentage of change to baseline over time. The increasing phenomena reveal the new normal life has been adapted. Fig. 2b shows weekly mean confirmed cases in Brazil that grew since the middle of April 2020. Note that the number of confirmed cases here has been normalized across all countries considered in this dataset. Fig. 2c shows that Malaysia’s human mobility reached the lowest activity of retail and recreation percent change to baseline in the middle of May 2020 and then gradually increased its percentage of change to baseline. The duration of low activities was around a month, starting from the middle of April to May 2020. After May, the increasing activities were recorded, revealing new normal life has been adapted. Fig. 2d shows weekly mean confirmed cases in Malaysia that grew starting from the middle of March 2020. However, starting from May 2020, the weekly mean confirmed cases were decreasing. Note that the number of confirmed cases here has also been normalized across all countries considered in this dataset. Fig. 2e shows that Indonesia’s human mobility reached the lowest activities percent change to baseline in the middle of May 2020 and then gradually increased its percentage of change to baseline. The increasing phenomena show that the new normal life has been adapted. Fig. 2f shows weekly mean confirmed cases in Indonesia that grew exponentially since the end of March 2020 as the number of tests increased. Note that the number of confirmed cases here also has been normalized across all countries considered in this dataset.

**Figure 2.**
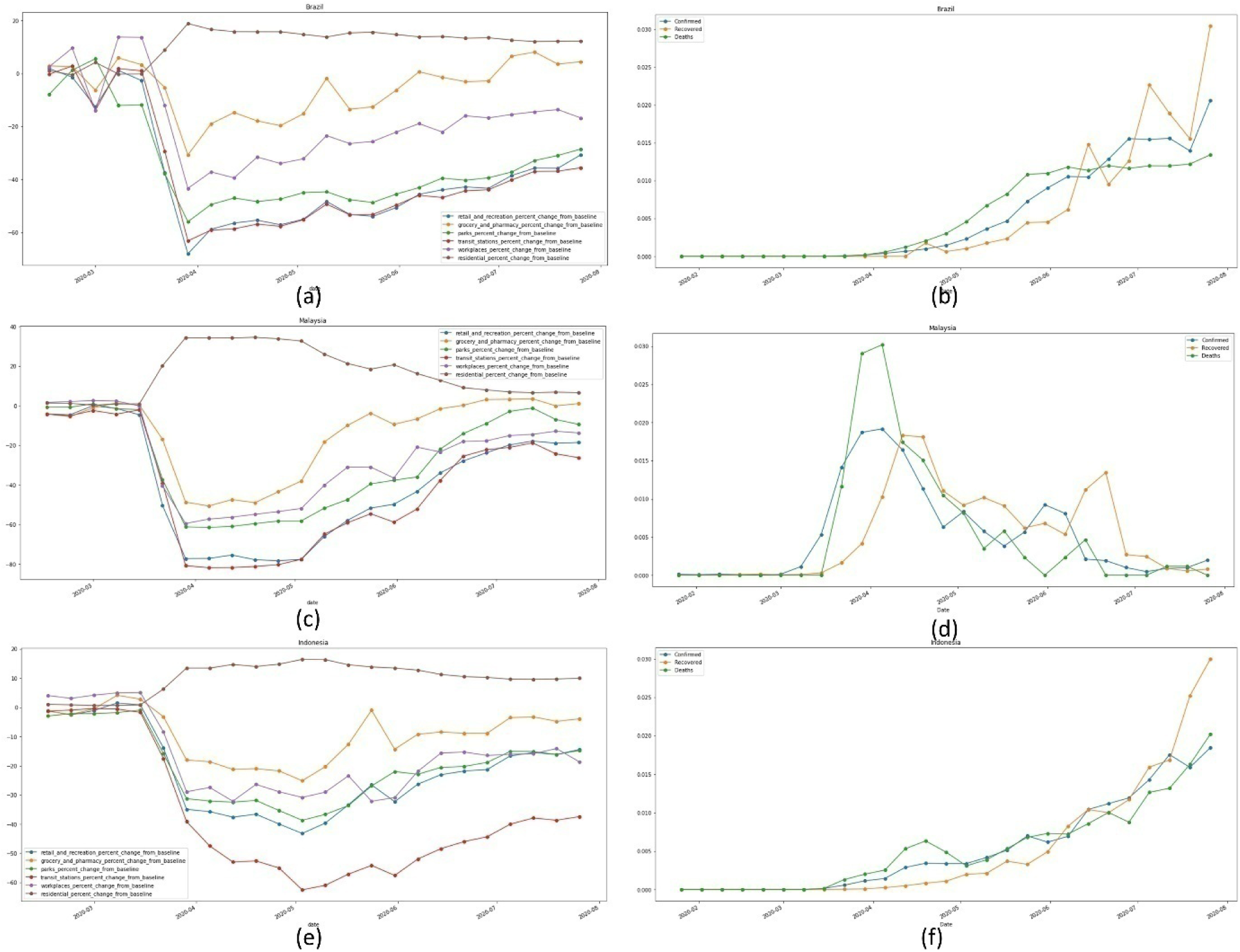
(a) Human mobility dynamics in Brazil (b) Weekly mean confirmed cases in Brazil (c) Human mobility dynamics in Malaysia (d) Weekly mean confirmed cases in Malaysia (e) Human mobility dynamics in Indonesia (f) Weekly mean confirmed cases in Indonesia

To answer the question of whether there is any correlation between the decreasing activities before new normal and weekly mean confirmed cases after the new normal, time-lagged cross correlation was carried out.

### 2.4. Statistical Analysis

The Jakarta region investigation in Fig. 3 showed a strong positive correlation when weekly mean confirmed cases time series are correlated with weekly mean human activities with an o set of −2. The weekly mean confirmed cases strongly relate to weekly mean human activities after around two weeks.

**Figure 3.**
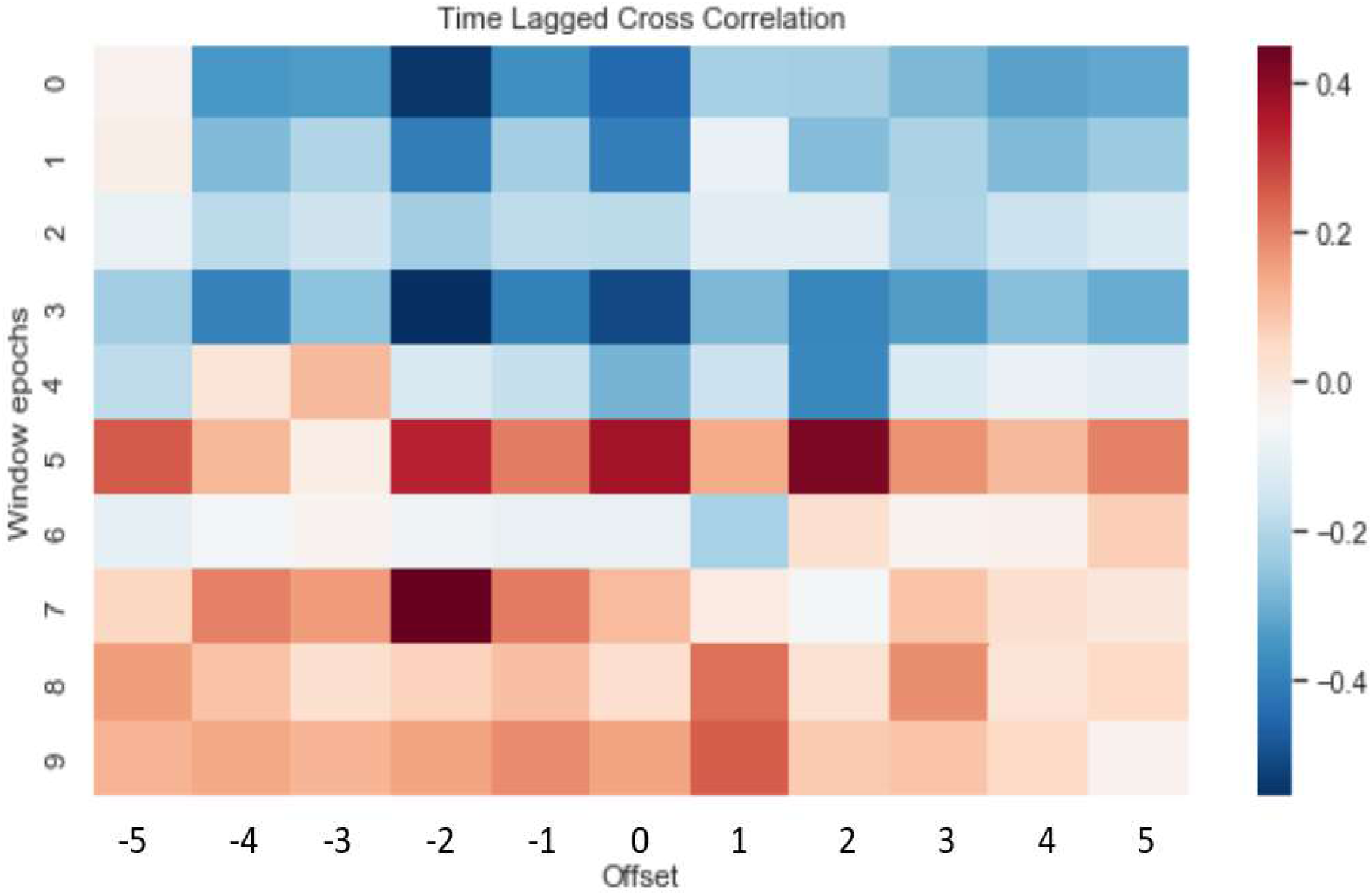
Time lagged cross-correlation between weekly mean confirmed cases and human activities in Jakarta region

Based on the investigation in Fig. 4, we make a correlation test of all countries between weekly mean confirmed, recovered, and death cases with weekly mean human activities with an offset of −2 (2 weeks before). It can be concluded that weekly mean confirmed cases are positively correlated to weekly mean workplaces and transit stations percent change from baseline by 0.42 and 0.43 with correlation p-value of 0.003 and 0.002, respectively. Weekly mean confirmed cases are negatively correlated to weekly mean residential percent change from baseline by −0.33 with a correlation p-value of 0.03. It means that weekly mean confirmed cases correlate to weekly mean workplaces, transit stations, and weekly mean residential percent change from baseline with statistically significant (p < 0.05).

**Figure 4.**
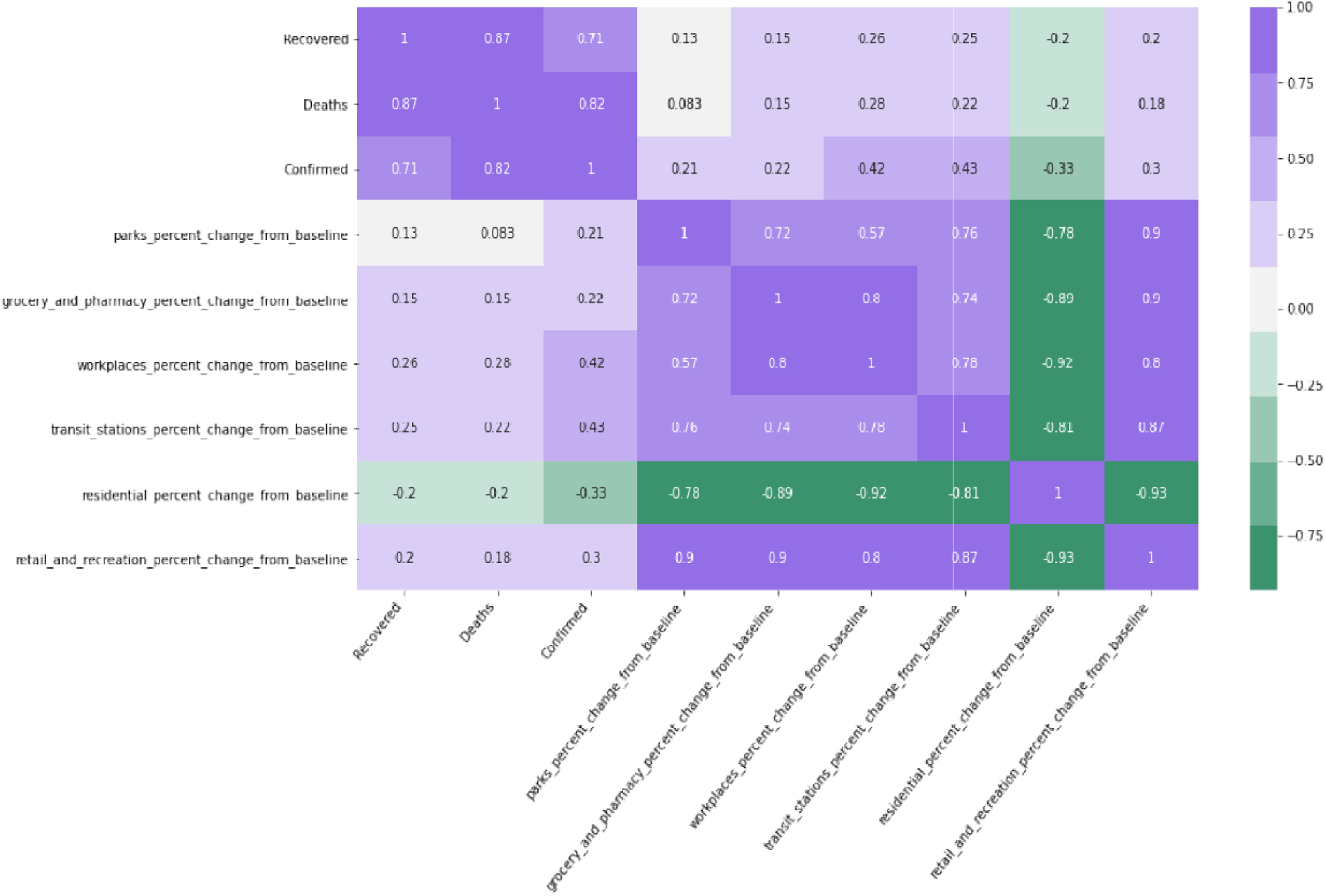
Pearson correlation of weekly mean confirmed, recovered, death cases and weekly mean human activities of all countries with o set of −2

### 2.5. Multivariate Time Series Analysis via Explainable Deep Neural Network

In this research, multivariate time series data set of 59 factors from 55 countries consists of 6 behavioral, 21 COVID-19, one government, two geographical, three morphological, two economics, five health, two health facilities, one education, and 16 environmental categories. Each factor is captured daily, except environmental factors (UVs) are represented by daily mean observations. A variant of DNNs then trains the features called Long Short Term Memory (LSTM) [22] to predict the outcome series of daily 59 factors over 174 days (2020-03-22 to 2020-09-11). All features in the X dataset are normalized feature-wise into Xscaled following this equation 1:

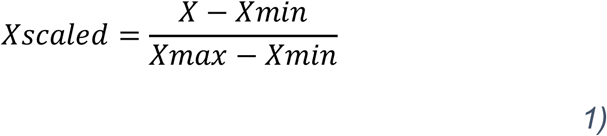

To achieve explainable prediction of Spatio-temporal data, we develop a Convolution-LSTM model that consists of 1 1D convolution layer and 2 LSTM layers with 59 hidden states followed by a fully connected layer with Sigmoid activation (Figure 4). The whole of LSTM units contains the input gate, the forget gate, and the output gate to capture Spatio-temporal correlation and dynamics of multivariate time-series data. The forward propragation flows from input input layer, hidden layers, and output layer followed by Sigmoid activation. After the learning phase, gradient-based optimization via backpropagation is utilized from which attribution maps (saliency maps) are generated. The visual attribution extracts attention to features that relevant to final Spatio-temporal time-series predictions (Fig. 5). Specifically, the method called GradCAM [23] is used to create its attribution maps. Grad-Cam is applied to the last hidden layer where its output activation is weighted with important weight associated with time-series predictions followed by Rectified Linear Unit (ReLU) activation.

**Figure 5.**
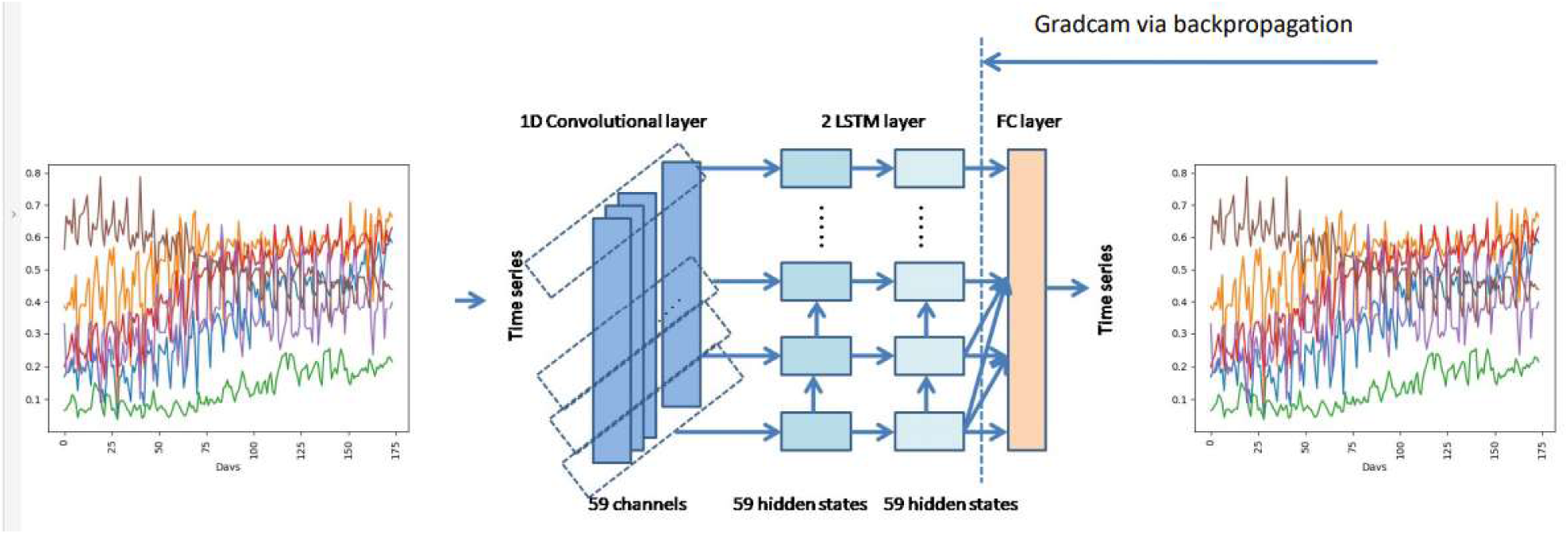
Convolution-LSTM architecture and its visual attribution via Gradcam

The formulation of Gradcam for LSTM hidden layer is given in equation 2 as

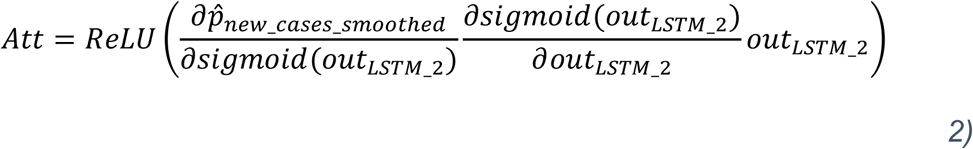

Where Att, 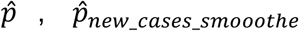 and *out*_*LSTM_2*_ are visual attribution, reconstruction output containing 59 factors, reconstruction output containing only new cases smoothed, and output of hidden layer of the second layer of LSTM (features), respectively. The gradients are obtained through partial derivatives and chain rule. The final visual attribution is transformed into clean visualization using ReLU activation by eliminating negative values.

## 3. Experimental Setup

We divide the dataset into training and validation, which are 55 and 4 countries, respectively. The total time-series length for all features is 174 days, 2020-03-22 to 2020-09-11. We use three architectures, which are 1D CNN with one layer, LSTM 1 layer, and proposed Convolution-LSTM (Conv-LSTM), which are validated to test the data set using Root Mean Squared Error (RMSE). The number of epoch is set to 3000. We use Adam optimizer to update weights for each iteration with a learning rate of 0.001. The best architecture based on best validation score is selected as a visual explanation model.

## 4. Results

Table 2 shows the (Root Mean Squared Error) RMSE to validate model architectures to predict the time-series outcome of Italy, Sweden, Indonesia, and Norway. The low RMSE indicates that the prediction accuracy of validation is high and impacts reliable feature attribution. After predictions, Spatio-temporal feature attention inside the Convolution-LSTM network can be visualized. High attention is represented by red color and gradually becomes blue as the attention value is decreased. The range of feature attention degrees is in the range of 0 to 1.

**Table 2.**
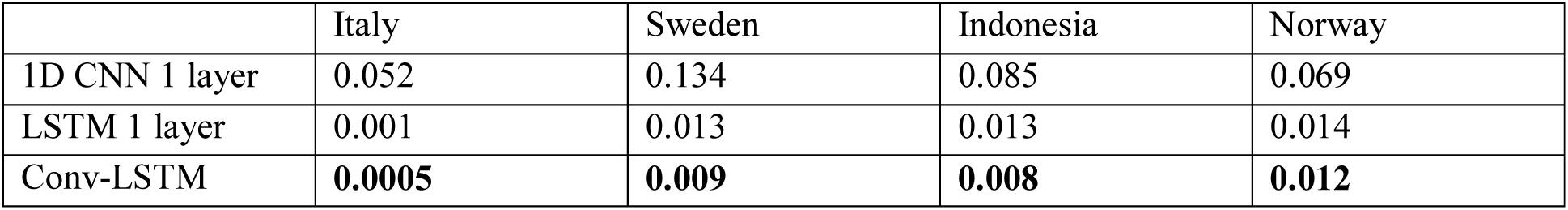
Root Mean Squared Error (RMSE) forLSTM on time-series prediction of Italy, Sweden, indonesia, and Norway.

Fig. 6 shows the reconstruction and actual of new cases smoothed in Indonesia. Even though the reconstruction result hardly matches the pattern of the actual one, the RMSE is 0.008, which is the lowest compared to other models (Table 2). Improper reconstruction is due to presumably Indonesia is a large country with heterogeneous character and behavior. Compared to Sweden, Norway, or Italy, Indonesia is greater in terms of population, and geographic area, leading to the need for more detailed and complete data in terms of region and period. Another suggestion is by adding more countries to be fed into the model; thus model can generalize well and reduce overfitting. For future works, data to be analyzed should be more detailed in terms of the region and the time-series sample period.

**Figure 6.**
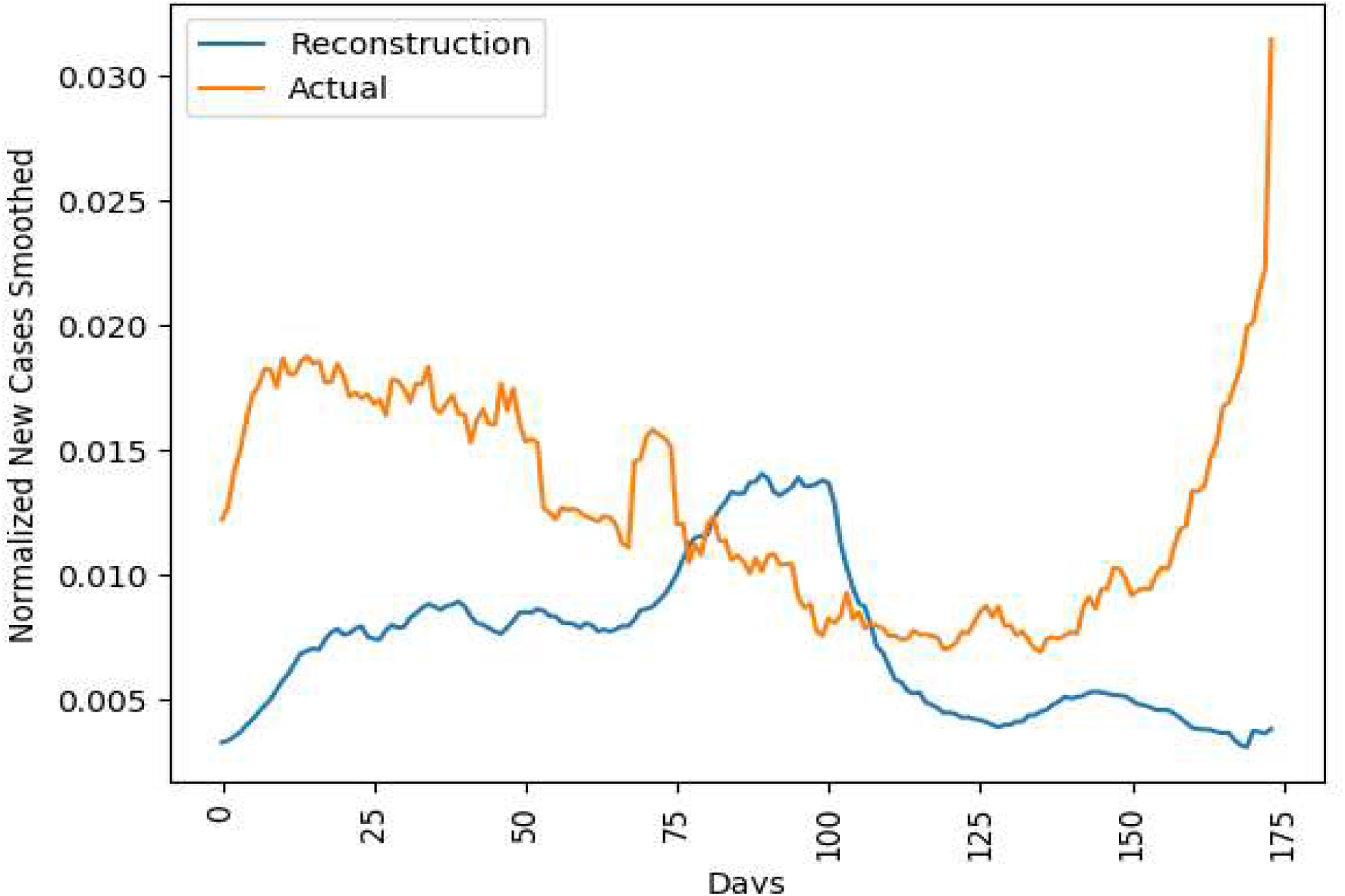
Actual and reconstruction of Indonesia

Fig. 7, 8, and 9 show actual and reconstruction of new Norway, Italy, and Sweden, respectively. They have a similar cases smoothed in time-series pattern between actual and reconstruction and thus have a better outcome than Indonesia. Coincidently, they are located in a similar geographical location of the northern subtropical area, which differs from Indonesia. It can be concluded that more samples are necessary to generalize countries that have a similar pattern to Indonesia’s COVID-19 case.

**Figure 7.**
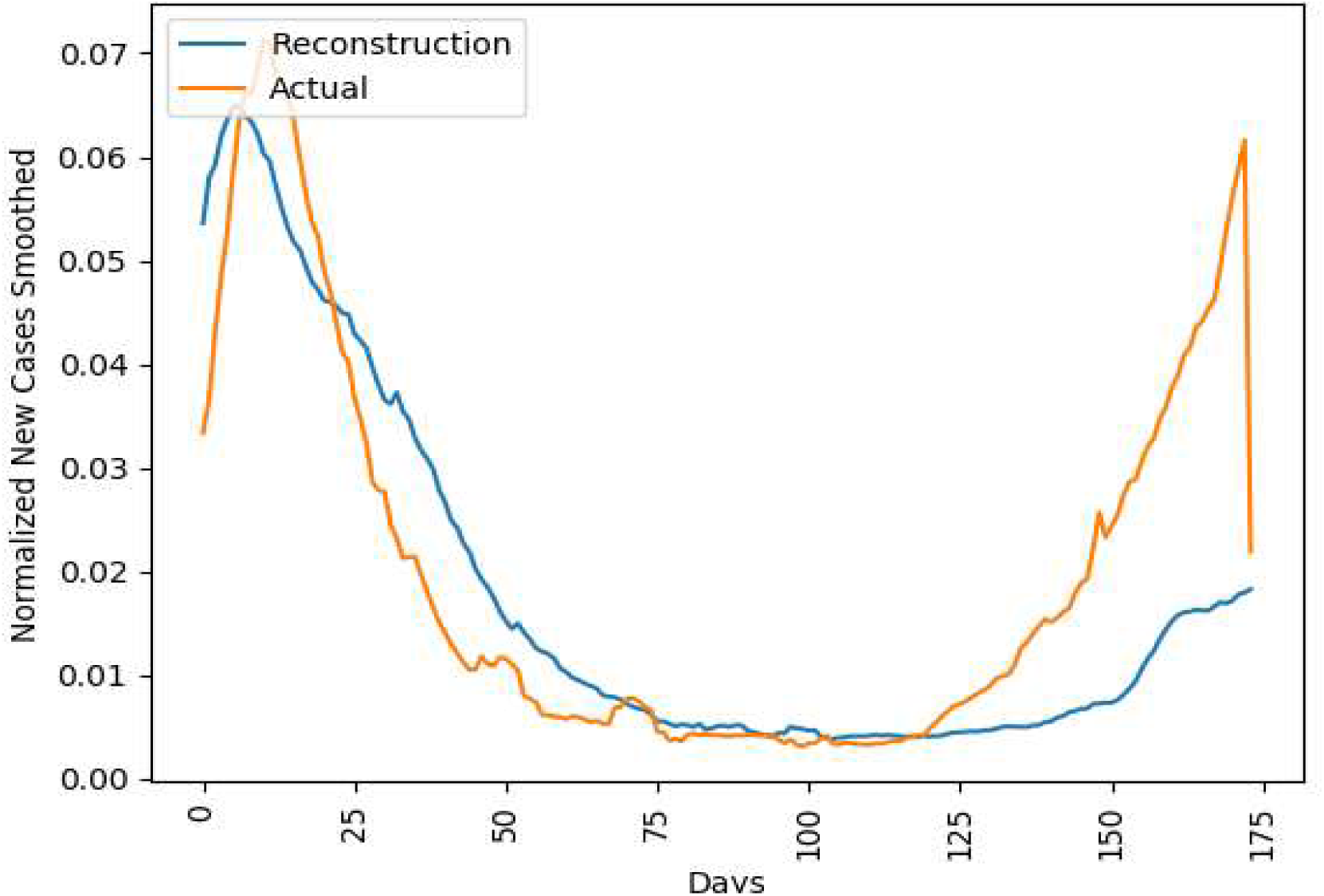
Actual and reconstruction of Norway

**Figure 8.**
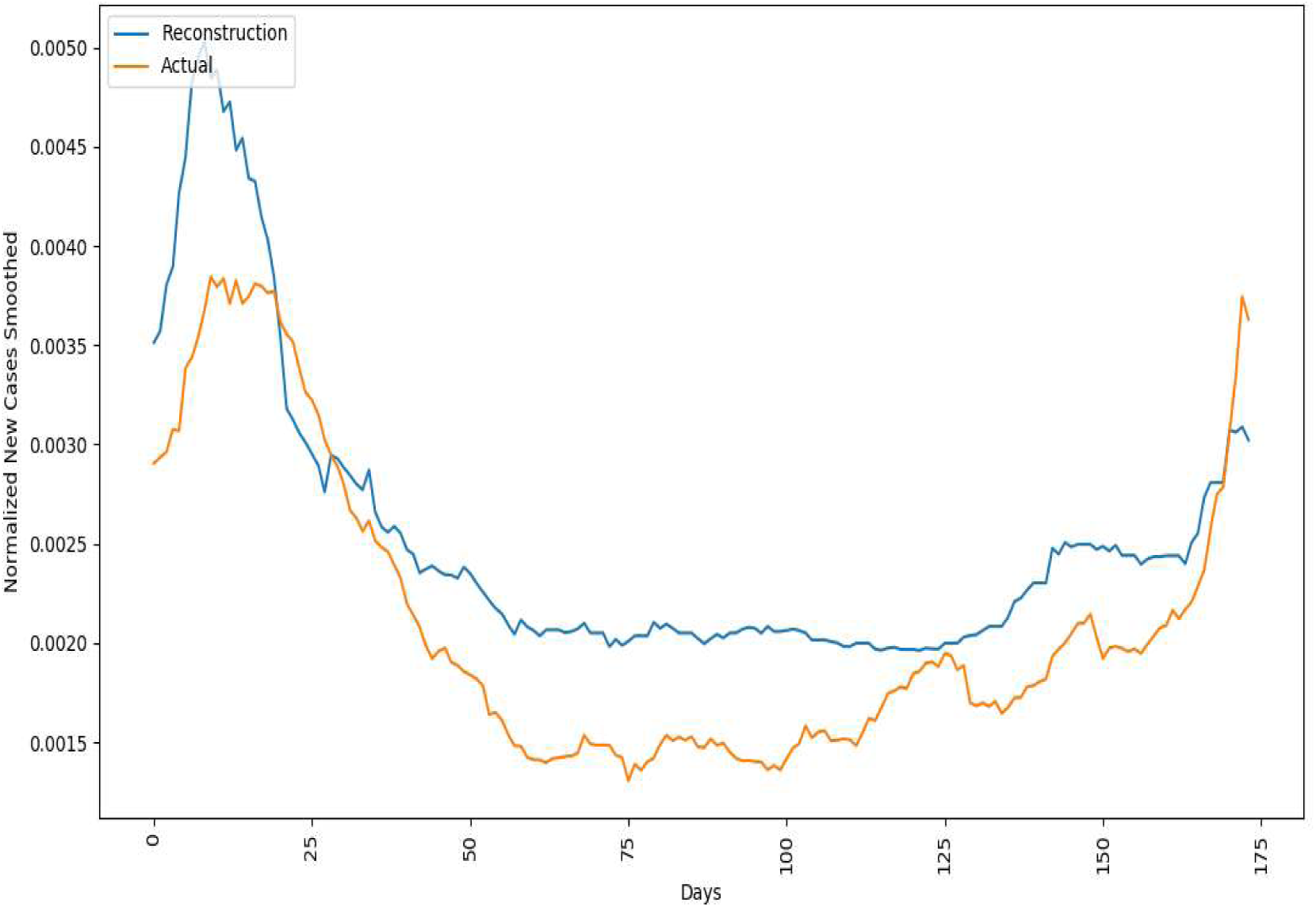
Actual and reconstruction of Italy

**Figure 9.**
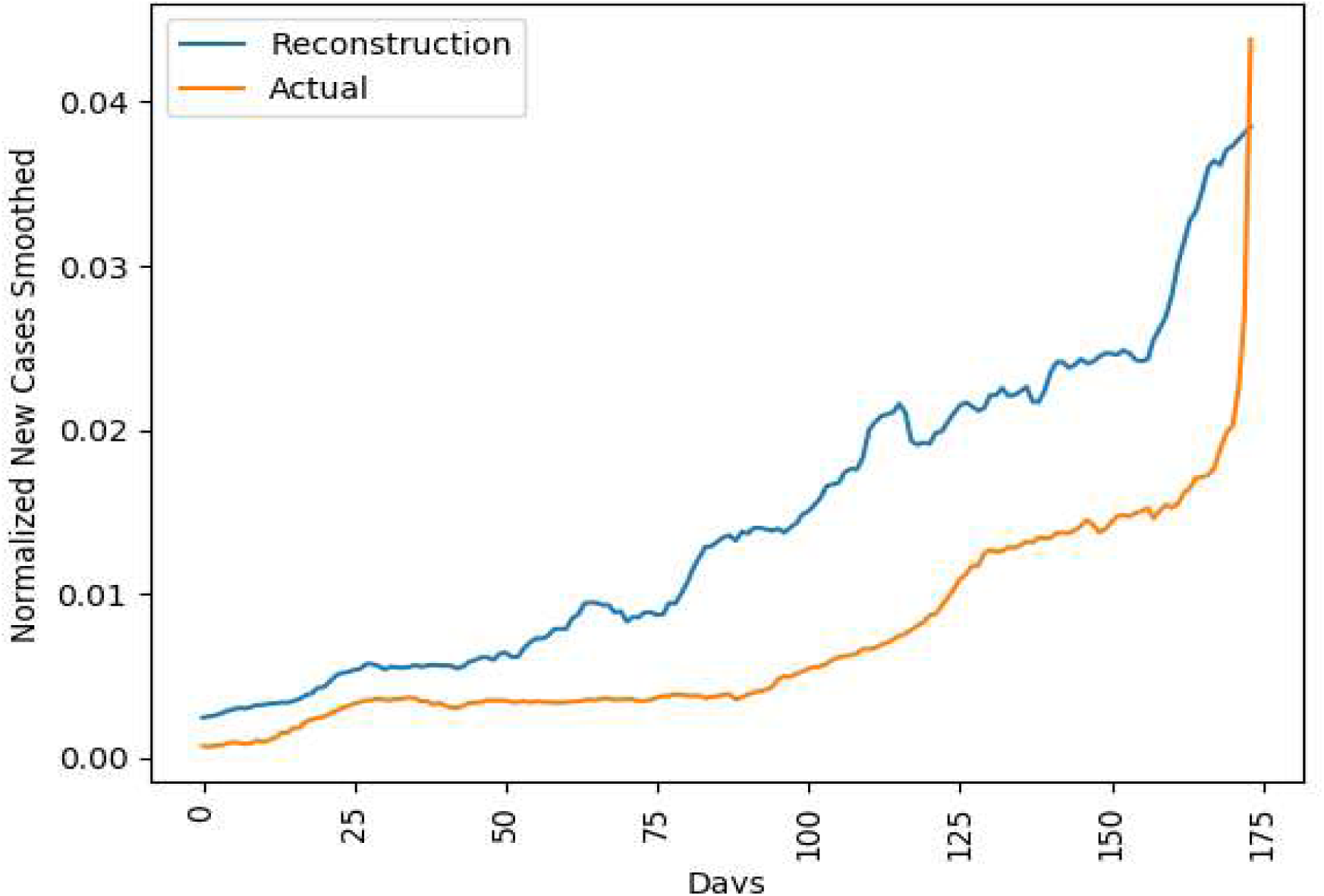
Actual and reconstruction of Sweden

Fig. 10 shows the example of the visual attribution of Conv-LSTM prediction on a series of daily new COVID-19 cases in Italy. When the visual explanation is investigated, network put high attention on the residential, retail and recreation percent change from baseline at the beginning of time. It regards changing activity on those area contributes to the COVID-19 daily new cases pattern over the time in Italy. Total test per thousand, number of test units, and environmental factors of UVDDC and UVDDCerr at the beginning of time also influence the new cases over time. The number of aged 70 people at the end of the time contributes to the pattern of new cases growth over time. The number of hospital beds per thousand is linked over time by a network which suggests an important factor to be considered. Factors that have been put high attention at the beginning and the end of the time are the number of new deaths over time and environmental factor of UVDDFerr. Over time new cases smoothed follows environmental factor of UVDDFerr pattern which decreases at the beginning of time and then increases again at the end of time. It indicates the influence of UV in affecting the graph of COVID-19 cases, especially in open spaces (the parks percent change from baseline influences new cases at the small amount at the beginning and the end of time) as the dynamics of UV in northern subtropical countries.

**Figure 10.**
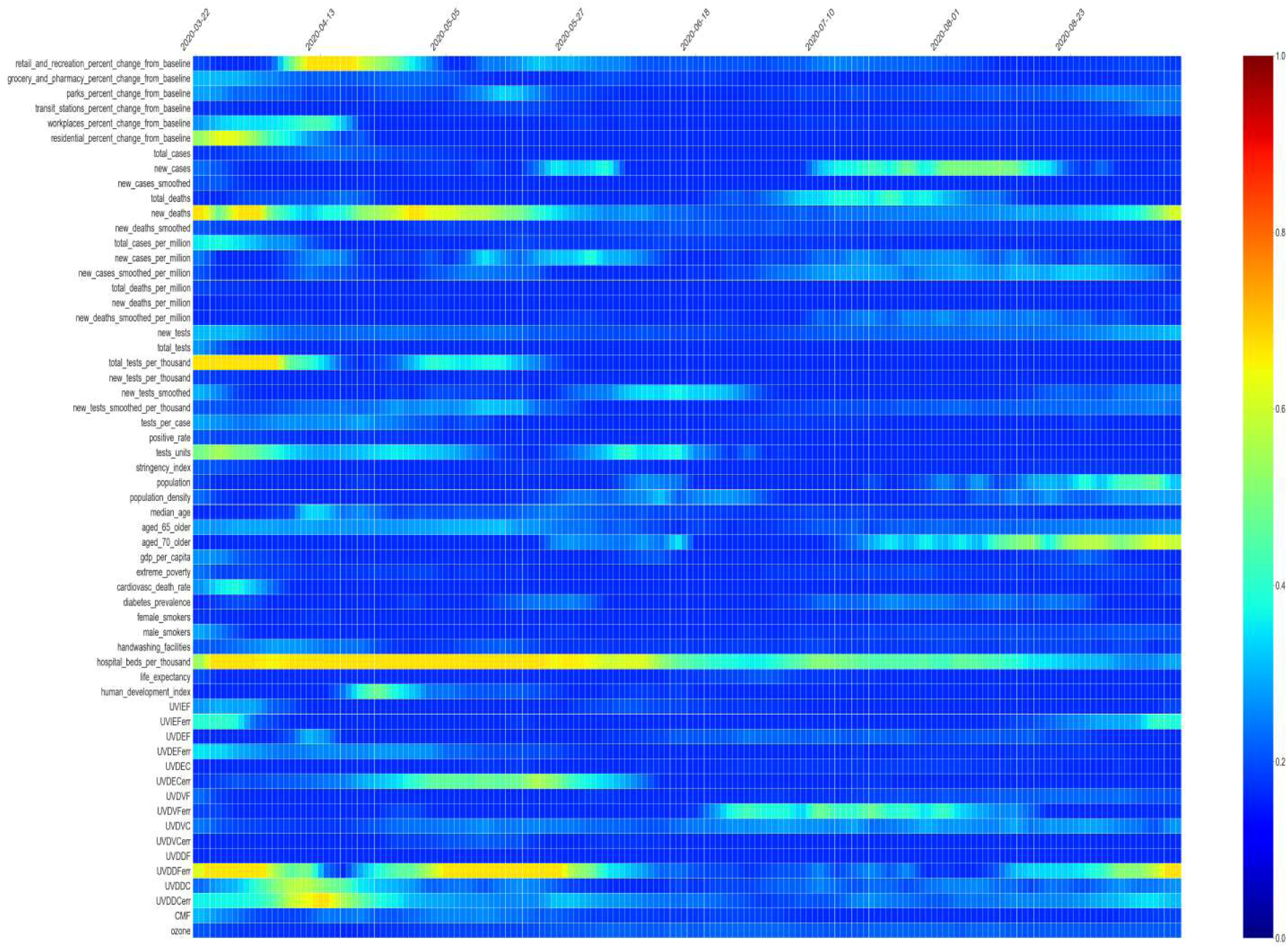
Time and feature attention corresponding to a prediction for new daily COVID-19 cases in Italy

Fig. 11 shows the visual attribution map of The Conv-LS M network that put attention into workplaces percent change from baseline almost the entirety of the time\ while insufficient attention on other places. The number of test units influences new cases initially while median age people contribute in the middle of time. Cardiovascular death rate also is attributed by the network at the beginning and the end of the time. Same as Italy, the number of hospital beds per thousand gives significant contribution at the entire time. Environmental factors of UVIEFerr, UVDEF, and UVDDF affects new cases smoothed time series pattern in the middle of the time.

**Figure 11.**
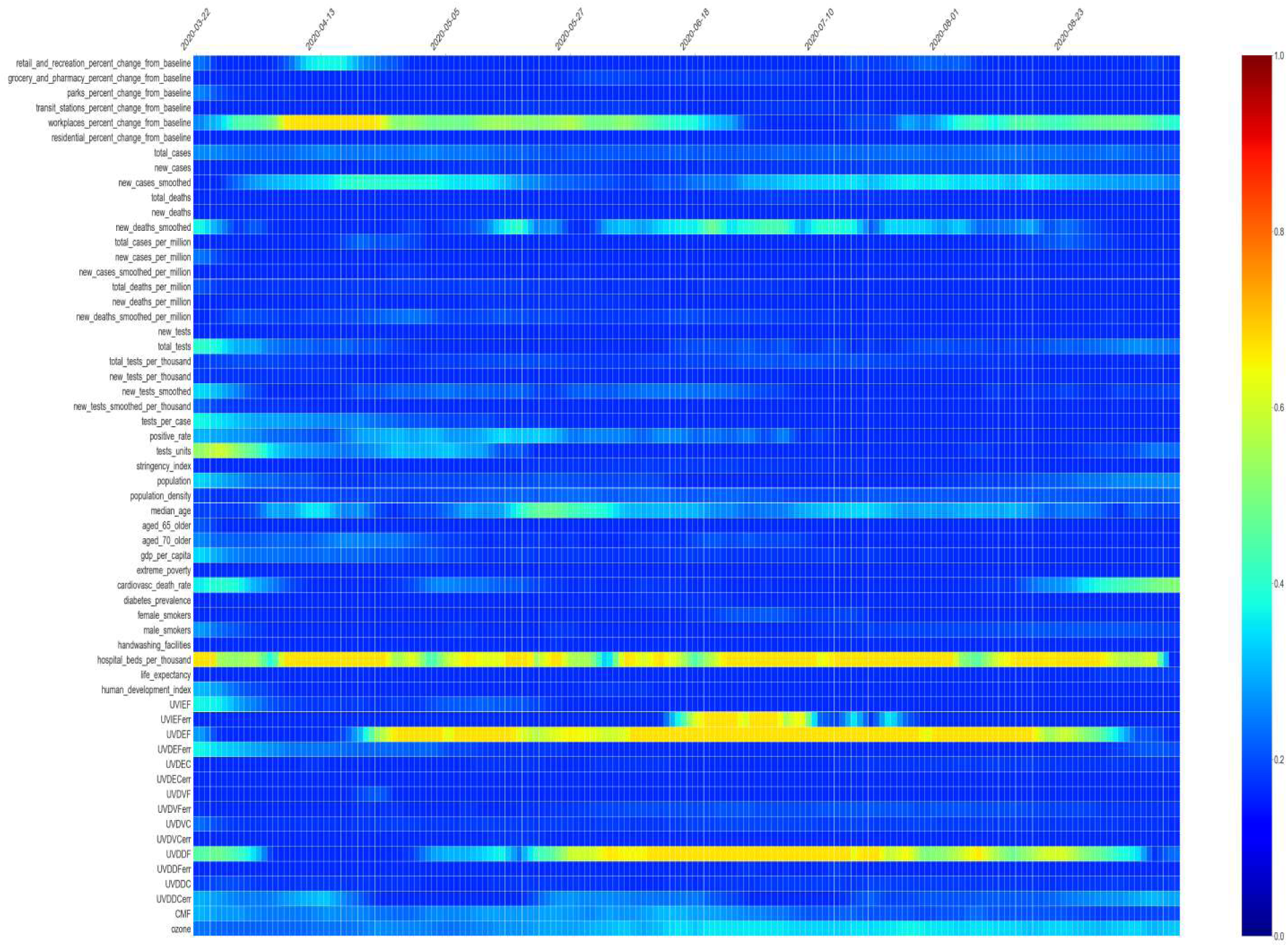
Time and feature attention corresponding to a prediction for new daily COVID-19 cases in Sweden

Temporally, attention to behavioral factors only highlights workplaces change to baseline over time. It regards open space as a park contributes to the COVID-19 daily new cases. UV index and dose increase over time in the northern subtropical countries toward summer. Correspondingly, the visual attribution also shows the degree of attention on environmental factors. It indicates the influence of UV on daily new COVID-19 cases, especially in open spaces like parks. The contribution of aged people is not as intense as Italy and Norway revealing Sweden’s success in separating aged people and younger ones.

Fig. 12 shows the example of the visual attribution map of Conv-LSTM prediction on series of daily new COVID-19 cases in Norway. Conv-LSTM network put attention into residential, grocery, and pharmacy percent change from the baseline while insufficient attention on transit station percent change from baseline. It regards closed space contributes to the COVID-19 daily new cases. In Norway, the difference between Norway and Sweden is th t closed space activities like in workplaces are not given as much attention as Sweden of contribution to daily new COVID-19 cases. It can be understood since the Sweden government’s treatment of society is not as strict as Norway [25]. Visual attribution map also shows that government stringency is one of the critical factors contributing to daily new COVID-19 cases. Consequently, as shown in Fig. 7 and 9, Norway and Sweden have different new cases smoothed pattern overtime where the former decreased and then increased while the latter is monotonically increasing. Besides, the number of test units, total tests per thousand, and the number of new tests initially affect the pattern of new cases smoothed over time.

**Figure 12.**
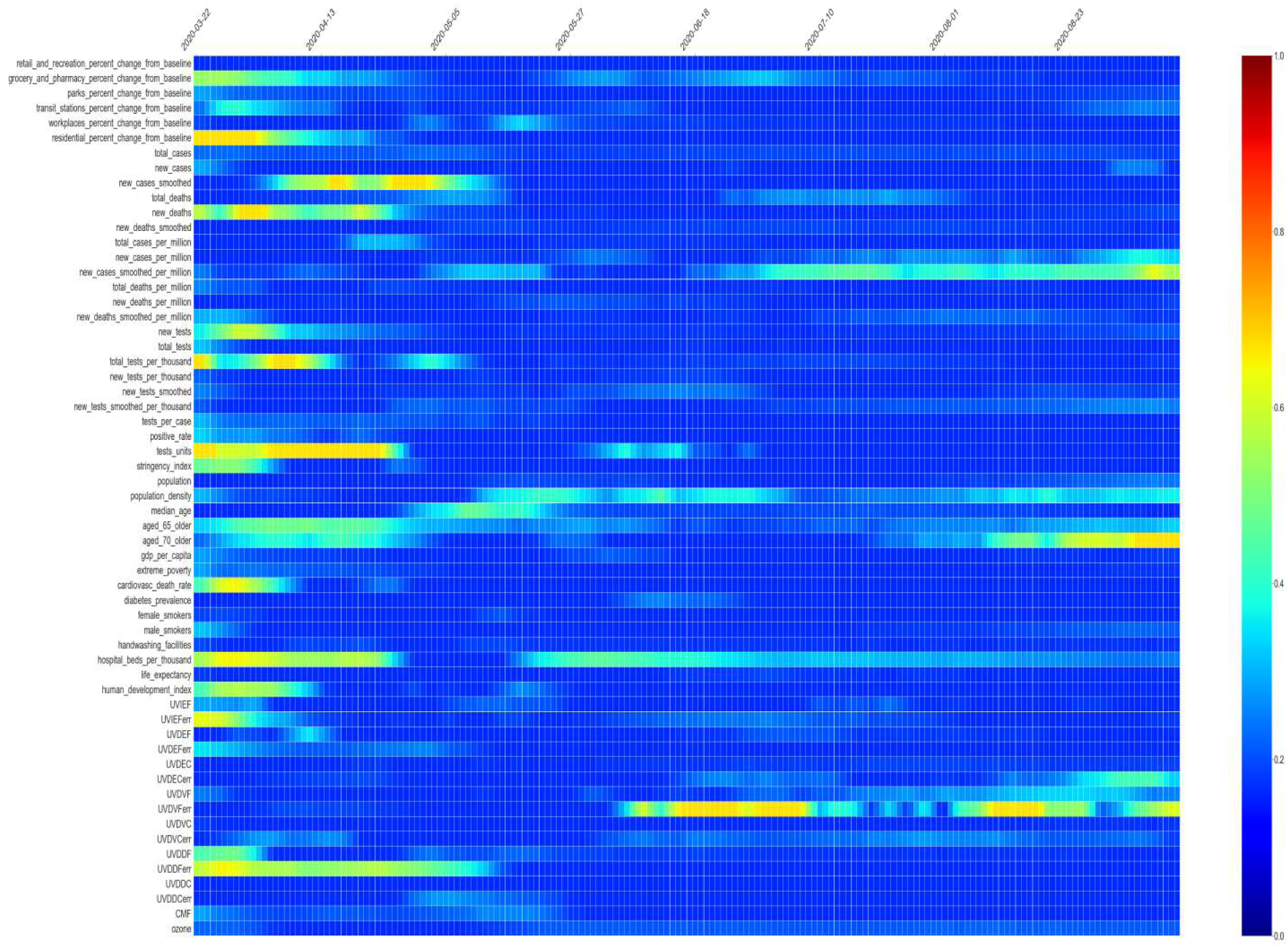
and feature attention corresponding to a prediction for new daily COVID-19 cases in Norway

The morphological factor of the number of aged 70 people at the end of time contributes to new cases smoothed. Human index and health factors of cardiovascular rate and number of hospital’s bed are also factors that cannot be underestimated. Environmental factors like UVIEFerr, UVDVFerr, and UVDDFerr contribute to the new cases smoothed in Norway.

Fig. 13 shows the example of the visual attribution map of Conv-LSTM prediction on a series of daily new COVID-19 cases in Indonesia. Unlike Italy, Sweden, and Norway, which are located in the northern subtropical region, the Conv-LSTM network puts high attention on behavioral factor of the residential percent change from baseline. It regards open spaces are safer than closed spaces like residential, grocery, and pharmacy where human-to-human transmission intensely occurs and open spaces like parks are not given much attention.

**Figure 13.**
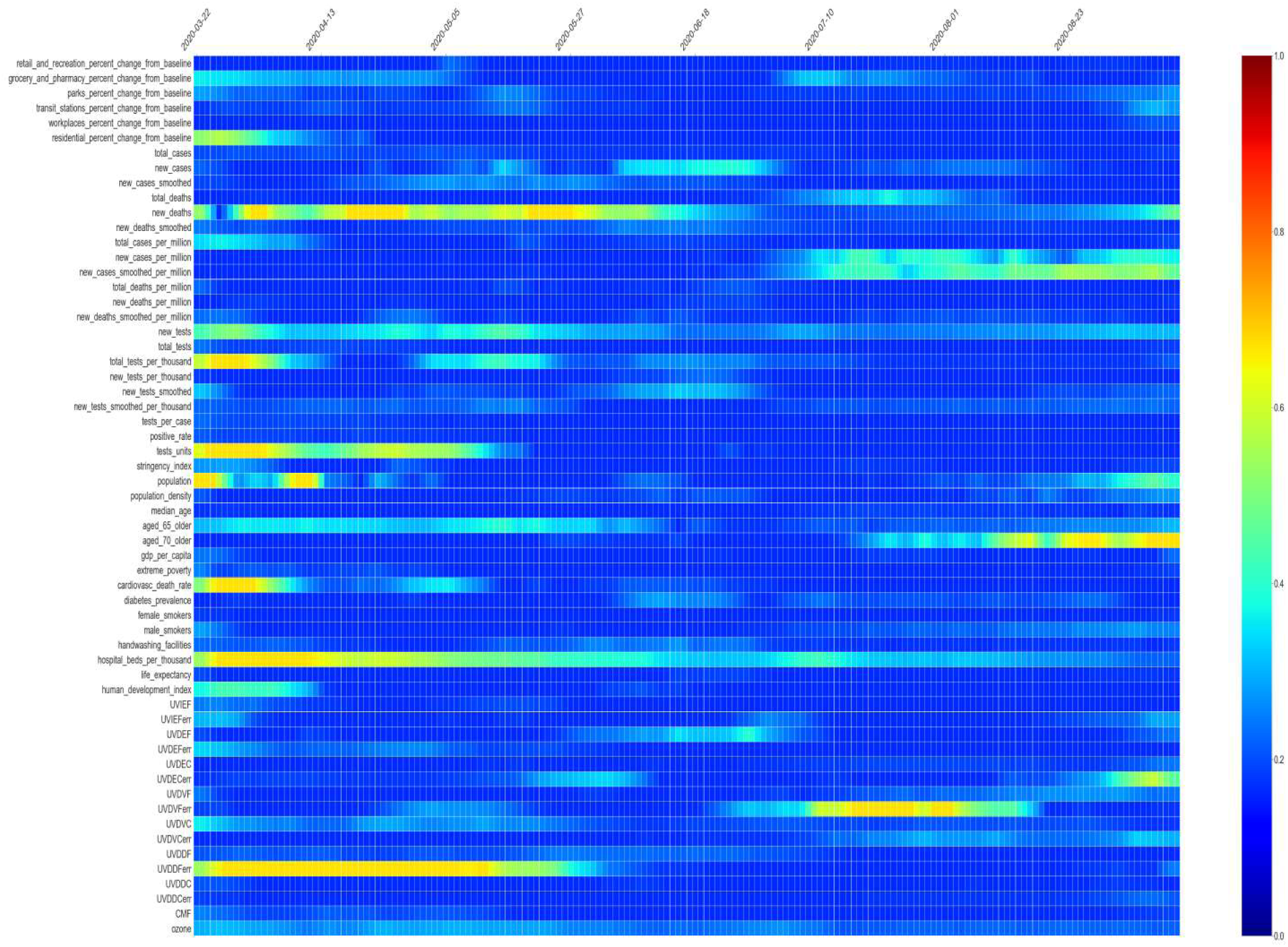
Time and feature attention corresponding to a prediction for new daily COVID-19 cases in Indonesia

Low degree of attention on activity in parks, grocery, and retail and UVIEF indicate that open spaces with low intensity of human-to-human interaction are helped by environmental factors such as UV (UVDDFerr at the beginning of time and UVDVFerr at the end of time). Indonesia’s daily new COVID-19 cases also depend heavily on the number of tests performed across the country at the beginning of time (number of new tests, total tests per thousand, and tests units). The big population also contributes to the new cases, especially if the number of tests grows over time. The number of hospital’s beds for the entire time is also an important factor contributing to new cases smoothed.

## 4. Conclusion

While environmental factor correlates to the global spread of COVID-19 pandemic, we believe that it is not just a standalone factor. Some other factors, like morphological and behavioral factors, influence the spread and growth of COVID-19 cases. Human activity also influences the spread and growth of COVID-19 cases via human-to-human transmission, especially in workplaces, residential, and groceries where direct human interaction is intense in closed spaces. Based on direct evidence, even though there is an indication that COVID-19 is seasonal flu where there is interchanging conditions between northern subtropical, tropical, and southern subtropical locations, some other anticipation should be taken into account:

1. The new normal life is an inevitable thing in daily life where wearing a mask, hand washing, increasing the hospital’s capacity, and minimizing the number of activities that make up the crowd should be concerned.
2. Open space is safer than in a closed room with a crowd due to the UV light and air circulation. Unfortunately, the victims mostly occur in closed spaces such as groceries, residential, and workplaces.
3. For tropical countries, an abundance of UV light helps withstand the COVID-19 spread, especially in open space. There must be a good balance between activities inside and outside rooms, especially at noon, where the level UV index is high. In closed spaces like workplaces, it is suggested to expose UV rays in the room before leaving the places when the work hour ends.
4. For subtropical country residents, wearing a mask is compulsory while living in an open space and during the cold season. By the time of the summer season, people in subtropical countries should take advantage of the high UV index level.

## Data Availability

Data set are publicly available on the internet

## Competing Interest

Authors have no competing interests to declare.

## Author Contributions

**NovantoYudistira**Conceptualization;Data curation; Formal

analysis;Methodology;Software; Visualization; Writing - original draft**Sutiman Bambang**

**Sumitro**: Conceptualization; Data curation; Formal analysis;

Supervision. **AlberthNahas**: Roles/Writing - original draft; Writing - review & editing;

Validation**Nelly Florida Riama:** Data curation; Investigation; Writing - original draft;

Writing - review & editing;Validation

## Notes

### Competing Interest Statement

The authors have declared no competing interest.

### Funding Statement

None

### Author Declarations

University of Brawijaya

